# Automated hippocampal sclerosis detection, using AID-HS, shows robust performance across multi-centre paired 7T and 3T MRI

**DOI:** 10.64898/2026.08.27.26356343

**Authors:** Cornelius Kronlage, Mathilde Ripart, Rory J Piper, Martin M Tisdall, David W Carmichael, Torsten Baldeweg, John S Duncan, Jonathan O’Muircheartaigh, Maria H Eriksson, Chiara Casella, Philippa Bridgen, Tobias Bauer, Simon R Bouschery, Annalena Lange, Eberhard D Pracht, Tony Stöcker, Rainer Surges, Theodor Rüber, Krzysztof Klodowski, Christopher T Rodgers, Thomas E Cope, Konrad Wagstyl, Sophie Adler

## Abstract

**Background:** Hippocampal sclerosis (HS) is a common cause of drug-resistant focal epilepsy (DRFE) and amenable to neurosurgical treatment. Detection relies on MRI but can be challenging. 7 Tesla (T) ultra-high field MRI and automated MRI post-processing tools have independently been shown to improve radiological diagnosis of HS. However, combining these approaches remains underexplored. This study evaluated whether AID-HS, a tool for HS detection developed using 3T MRI, generalises to 7T MRI data.

**Methods:** We collated a dataset of paired 3T and 7T T1-weighted MRI from four epilepsy centres, including 23 patients with HS, 39 healthy controls, and 23 individuals with focal cortical dysplasia as disease controls. Histopathology served as the gold standard for defining HS where available (n=7), otherwise radiological findings (n=16). AID-HS was applied to images acquired at both field strengths, and sensitivity and specificity for detection and lateralisation of HS were compared. Additionally, agreement of hippocampal features across 3T and 7T was evaluated.

**Results:** We found no evidence of a difference in performance of AID-HS between 3T and 7T. Sensitivity for detection of unilateral HS was 63% (12/19) at 3T and 68% (13/19) at 7T (McNemar’s exact test p=1.0). Specificity in controls was 97% (60/62) at 3T and 100% (62/62) at 7T (p=0.5). Bilateral HS was correctly flagged in 3 of 4 cases using feature-based criteria, with high specificity in controls. Quantitative hippocampal features showed moderate to good agreement across field strengths (ICC 0.70 to 0.98), with small differences observed for volume and thickness estimates.

**Conclusion:** AID-HS provides robust detection and lateralisation of HS across multiple 7T MRI centres, highlighting its potential to enhance lesion detection. Future work is needed to investigate whether models trained on 7T data can leverage the improved image quality for further gains in HS detection performance.

## Introduction

Hippocampal sclerosis (HS) is one of the most frequent lesions underlying drug-resistant focal epilepsy (DRFE) and the most common pathology targeted by epilepsy surgery [Blumcke et al., 2017]. Randomised controlled trials have shown that temporal resections for HS are highly effective in achieving seizure control compared to medical treatment alone [Jobst and Cascino, 2015]. HS manifests with epilepsy in children and adults and is considered to have various genetic, developmental and acquired aetiologies [Deleu et al., 2026].

Clinical detection of HS relies on magnetic resonance imaging (MRI), with three main radiological characteristics described: decreased volume, disruption of internal structure and increased T2w signal of the hippocampus [Blümcke et al., 2013]. However, HS can be subtle and escape visual detection on MRI; it has been reported on histopathology in a significant portion of resections performed in MRI-negative patients (10% in [Bien et al., 2009], 9% in [Wang et al., 2013]). The pre-surgical detection of HS on MRI is associated with favourable post-surgical outcomes [Jones and Cascino, 2016]. Accurately identifying HS in DRFE can enable more patients to be considered for epilepsy surgery, may reduce the need for additional investigations such as invasive EEG or positron emission tomography (PET), and shorten delays that could worsen outcomes [Janszky et al., 2005] as hippocampal atrophy progresses over time [Wu et al., 2026].

This clinical need for improved identification of HS has motivated the development of computational tools for analysing MRI images. Some tools first extract radiologically meaningful features (such as hippocampal volume or T2/FLAIR intensity) and then use a statistical or machine learning procedure to derive a prediction [Belke et al., 2025; Caldairou et al., 2021; Mo et al., 2019; Ripart et al., 2024a]. Others apply deep learning techniques such as convolutional neural networks (CNN), which are trained end-to-end with MRI data as inputs and diagnostic labels as outputs [Chang et al., 2023; Ito et al., 2021; Kim et al., 2022]. Detection accuracies range between 88% and 95%; however, a direct comparison of diagnostic performance of different models is not straightforward because separate cohorts and different evaluation metrics are used in many cases.

Research efforts have also focused on how to acquire more informative MRI data for identifying epileptogenic lesions in DRFE. Notably, ultra-high field MRI (using static magnetic field strengths of 7T or higher) provides increased signal-to-noise ratio and increased spatial resolution, although there are technological obstacles such as radiofrequency field inhomogeneities [Ladd et al., 2018; Springer et al., 2016]. Studies applying 7T MRI in people with focal epilepsies have shown identification of new relevant findings in 20-30% of cases [van Lanen et al., 2021], with HS accounting for a proportion of the additional findings in most studies (i.e., 1/3 [Bubrick et al., 2022], 3/18 [Burkett et al., 2026], 0/4 [Chen et al., 2021], 1/9 [Klodowski et al., 2025], 2/9 [van Lanen et al., 2025], 0/6 [Vecchiato et al., 2025]).

While automated lesion detection and 7T imaging have independently been shown to increase diagnostic yield, their combination remains underexplored. Machine learning models rely on training data being representative of possible inputs used for inference. A mismatch - in this case between 3T and 7T MRI - is termed a ‘domain gap’ or ‘domain shift’ and can result in degraded performance [Hoffmann, 2025]. At the same time, high-performing models typically require large training datasets. In comparison to the scale of clinical 3T cohorts [Gill et al., 2021; Ripart et al., 2024b; Ripart et al., 2025; Taylor et al., 2025], focal epilepsy 7T MRI studies are small, and clinical or histopathological confirmation for annotation of lesions is scarce. These factors currently limit the development of dedicated models using 7T MRI data. Consequently, it is desirable to apply models developed with large 3T MRI cohorts to 7T data both for interim use but also to serve as a baseline for future 7T specific prediction models, when more data becomes available.

Beyond robustness to domain shift, given the 20-30% diagnostic gain associated with visual assessment of 7T relative to 3T MRI, one might also expect similar benefits for automated analyses of 7T data. However, because epileptogenic lesions newly diagnosed at 7T are often visible on 3T MRI upon retrospective re-review [van Lanen et al., 2025; Vecchiato et al., 2025; Wang et al., 2020], it is conceivable that automated methods already capture subtle imaging features at 3T and therefore gain less from the enhanced image quality of 7T MRI.

AID-HS [Ripart et al., 2024a] is a recently published research tool for HS detection that was developed using 3T MRI of exclusively histologically verified HS patients, achieved state-of-the-art performance on a large multi-centre dataset, and has been independently validated on a clinical 3T cohort [Belke et al., 2025]. It uses the software HippUnfold [DeKraker et al., 2022] to extract five interpretable hippocampal features per hippocampus (volume, thickness, gyrification, mean and intrinsic curvature). Then a logistic regression classifier predicts whether and on which side HS is present based on these features. As HippUnfold was trained with sub-millimetric 3T data and tested on 7T data, we hypothesised that AID-HS would be robust to the domain shift between its 3T training data and 7T MRI.

In this study we collated a multi-centre dataset with paired 3T and 7T T1-weighted MRI from four different sites, spanning paediatric and adult age ranges, and selected patients with HS as well as healthy controls and disease controls. We used this dataset to compare the detection performance of AID-HS and its quantitative radiological features obtained at 7T *vs* 3T. See Figure 1 for an overview of the study.

**Figure 1:**
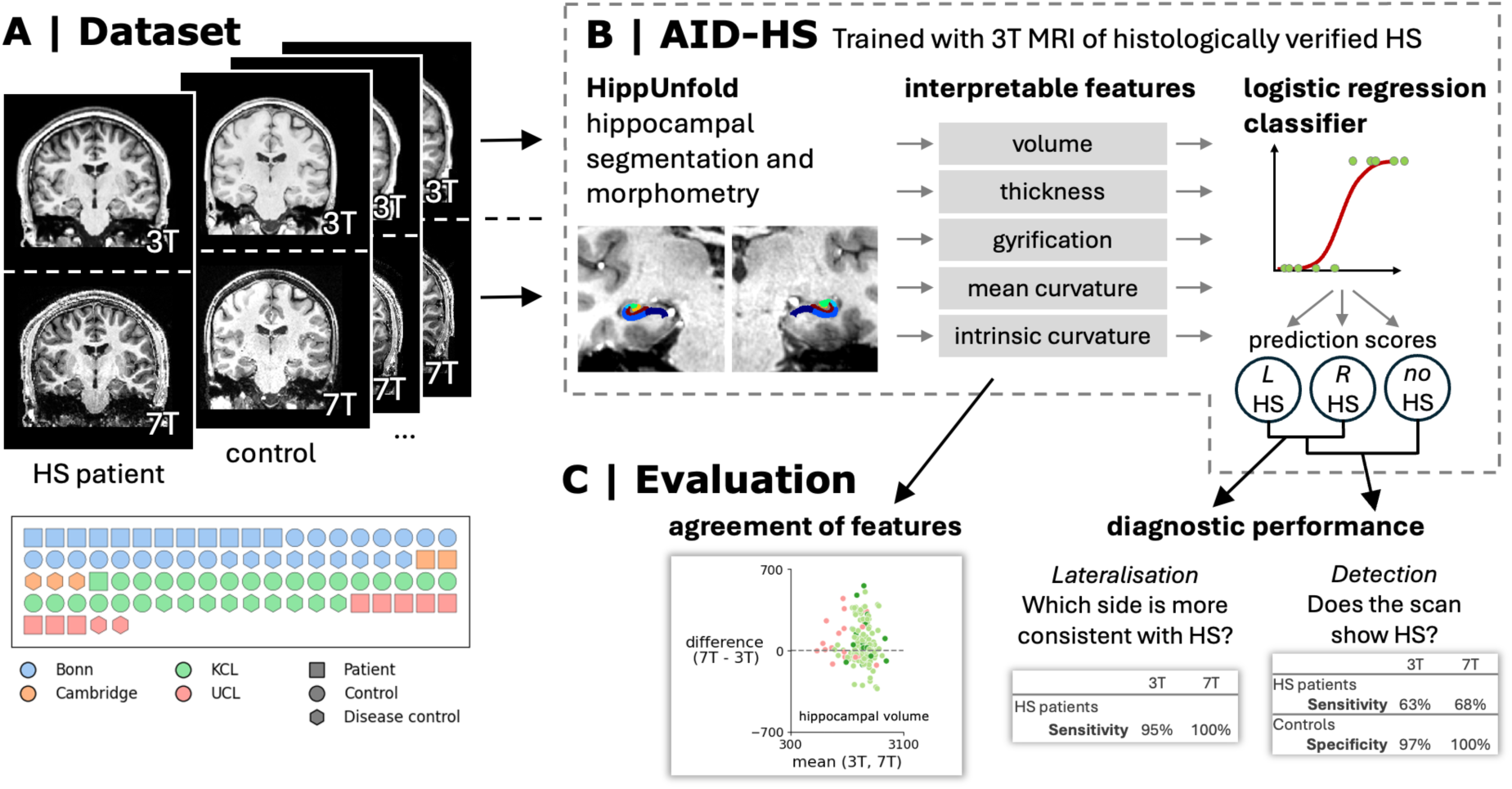
Study overview. (A) Paired 3T and 7T T1-weighted MRI volumes of 23 HS patients, 39 healthy controls and 23 disease controls from four tertiary care epilepsy centers were collated. (B) AID-HS was applied to 3T and 7T data separately. AID-HS includes hippocampal segmentation and feature extraction using the software HippUnfold. Extracted features are inputs to a logistic regression classifier that outputs scores for left HS, right HS and no HS. (C) The agreement of extracted morphological features as well as the overall diagnostic performance with respect to lateralisation and detection of HS was compared between 3T and 7T data.

## Methods

### Dataset

We collated 7T and paired 3T MRI data from four different epilepsy centres: University College London, UK (UCL); King’s College London, UK (KCL) [Vecchiato et al., 2025]; University of Cambridge, UK [Klodowski et al., 2025]; and University Hospital Bonn, Germany. At the respective sites, patients with drug-resistant focal epilepsy had undergone 7T MRI for clinical or research purposes. Ethical approval for data acquisition at each centre was as follows: Cambridge: UK National Research Ethics Service (23/WM/0008). Bonn: Ethics Committee at the Medical Faculty of Bonn (2020-83_3-BO, 2025-55-BO). KCL: UK Health Research Authority (IRAS 243811), UK National Research Ethics Office (ref. 18/LO/1766). UCL: Bradford Leeds Research Ethics Committee (REC ref: 22/YH/0171), King’s College London Research Ethics Committee (Development of Novel Magnetic Resonance Techniques Using Healthy Volunteers Study, KCL REMAS ref: 8700). The analysis was approved by the UK Health Research Authority (IRAS 301863).

For the present study, we defined three groups: patients with hippocampal sclerosis (HS), healthy controls, and patients with focal cortical dysplasias (FCD) as disease controls. Inclusion criteria for HS were: diagnosis of DRFE and radiologically identified HS on either 3T or 7T MRI, with multidisciplinary team consensus, or histopathologically verified HS. For FCD: diagnosis of DRFE and radiologically or histopathologically diagnosed FCD. For healthy controls: no neurological diagnosis and 7T MRI acquired with the same protocol as patients. For all groups, inclusion criteria included availability of both 7T and 3T T1-weighted MRI images. Participants with marked artefacts, e.g., due to movement, following manual image quality control were excluded.

As demographic and clinical variables, we included: age, sex, radiological findings (HS vs. unspecific/unremarkable) on 3T MRI and 7T MRI, whether surgery was performed, histology, and favourable outcome (defined as Engel or ILAE class I at least one year after surgery).

### Processing

We used high-resolution 3D T1-weighted images for analysis (Supplementary Table 1). AID-HS version 1.1.0 [Ripart et al., 2024a], which is based on HippUnfold version 1.1.0 [DeKraker et al., 2022], was used to generate automated hippocampal segmentations, extract volumetric and surface-based features and predict the presence and lateralisation of HS. AID-HS is packaged as a software pipeline that requires a T1-weighted MRI volume and demographic data (age, sex) as inputs, and generates quantitative segmentations, feature measurements, predictions and a summary PDF report as outputs without manual intervention.

We manually checked HippUnfold outputs with a segmentation QC score below 0.7 (a threshold suggested in the original publication [DeKraker et al., 2022]).

In AID-HS, scanner-specific effects can be compensated for by harmonisation of features with ComBat [Fortin et al., 2018] using a new site’s control subjects. In this study, for some sites, no paired 3T and 7T healthy controls were available. Furthermore, for some sites, 3T data had been acquired on multiple different scanners. Hence, we did not use harmonisation for the main analysis, but still show results obtained with ComBat harmonisation using all cohorts with paired healthy controls (Supplementary material).

To assess a possible laterality bias of the pipeline, we processed images flipped along the right-left axis and flipped the final output predictions back before evaluation.

Although AID-HS was developed with T1-weighted images, we also assessed whether high in-plane resolution T2-weighted images would yield higher performance. We processed all cases with both 3T and 7T T2-weighted images available with a modified version of AID-HS that uses the T2 modality-specific setting of HippUnfold.

### Visualisation

For a side-by-side visualisation of 3T and 7T data, corresponding volumes were co-registered using FreeSurfer (version 8.1.0) mri_coreg [Fischl, 2012] after skullstripping with mri_synthstrip [Hoopes et al., 2022]. 3T data and segmentations were realigned to the 7T space with trilinear interpolation for MRI data and nearest neighbor interpolation for segmentations.

### Performance evaluation and statistical analysis

To assess whether the distribution of males and females as well as age was similar in patients, controls and disease controls, we computed a Chi-Square test and, respectively, a Kruskal-Wallis test with post-hoc pairwise Mann-Whitney-U-tests with Bonferroni correction. For the Bonn dataset, as ages were provided in ranges spanning five years for anonymization purposes, values were imputed as the mean over the respective range (e.g., 18 for range 16-20) for statistical analysis. AID-HS classifier outputs can be evaluated with regards to correctly distinguishing patients from controls (detection) and correctly identifying the pathological side in HS patients (lateralisation). As summary measures of performance, we calculated sensitivity (fraction of correct predictions in HS patients) for detection and lateralisation, and specificity (fraction of correct predictions in controls) for detection. For comparing these performance metrics between paired samples of 7T and 3T data, we computed McNemar tests.

AID-HS uses a logistic regression classifier that predicts one of three classes: an asymmetry of hippocampal features consistent with left HS, right HS, or no asymmetry. It was not developed for the identification of cases with bilateral HS. However, it is possible to compare the values of hippocampal features to a normative control cohort modeled by a generalized additive model (GAM). In line with the original publication, we defined a threshold of any three or more outlier features consistent with HS (above/below the 5th or 95th percentile) bilaterally, out of a total of five features, as indicative of bilateral HS. We evaluated the sensitivity of this criterion in bilateral HS cases and its specificity in healthy and disease controls.

Hippocampal features and AID-HS prediction scores were compared visually in Bland-Altmann plots (mean vs. difference of paired measurements between 3T and 7T). For each feature, we fitted a mixed-effects linear regression model with subject as random effect, group (healthy control, disease control, patient) and hemisphere (ipsilateral or contralateral to HS, randomly assigned for controls) and their interaction as well as field strength (categorical levels: 3T, 7T) as fixed effects. For field strength as the covariate of interest, we report t-test p-values with Benjamini-Hochberg false discovery rate (FDR) correction. We also computed the intraclass correlation coefficient for absolute agreement for a single rater (ICC(A,1)) with field strength as ‘rater’ variable, using the python library pingouin [Vallat, 2018].

To assess whether model performance depends on the degree of hippocampal atrophy, we compared volume asymmetry indices in the groups of correctly detected vs. not detected HS patients (Mann-Whitney-U tests).

As a comparison for HippUnfold segmentations, we also applied SynthSeg, a widely used segmentation software that is tailored for generalisability through its domain randomization approach [Billot et al., 2023]. We obtained hippocampal volumes from the same T1-weighted inputs that were used for AID-HS and computed volume asymmetry indices as AI = 2 (vol_L_ - vol_R_) / (vol_L_ + vol_R_) [Ripart et al., 2024a]. To assess the diagnostic performance of Hippunfold and SynthSeg, we computed areas under the receiver operating characteristic curve (ROC AUC) for the binary classification of subjects as right HS vs. no or left HS, and left HS vs. no or right HS, based on volume asymmetry indices. For each condition, we report a weighted average (by number of left and right HS cases). 95% confidence intervals were obtained by bootstrapping (random resampling with replacement; 10,000 iterations).

For all tests, we set a significance threshold of p < 0.05.

### Data and code availability

The MRI data cannot be shared publicly due to privacy concerns. The code used for analysis is available at https://github.com/ckronlage/aidhs_uhf_pub/.

## Results

### Demographic and clinical data

Overall, the available dataset comprised 85 subjects based on available clinical diagnoses: 23 patients with HS, 39 healthy controls and 23 patients with FCD as disease controls. Data from the four sites included paired MRI of an additional 150 patients with DRFE with pathologies other than HS or FCD, or no known lesion. These MRI scans were not included in this analysis.

For key clinical data, see Table 1. Subjects had a median age of 23 years (range 8-53 years). Age distribution was different in the three groups; the median age of HS patients was higher than that of healthy controls (28 vs. 15 years; p=0.002). There were 49 male and 36 female subjects. There was no statistical relationship between group and sex (p=0.76). HS was defined by histopathological results for 7 cases (four of which were 3T MRI-negative and two 3T and 7T MRI-negative), and based on radiological diagnosis for 16 patients. Left HS accounted for more than half of the unilateral HS cases (15/19, 79%, 95% CI 56.7-91.5%).

**Table 1:** Multi-centre paired 7T and 3T MRI focal epilepsy cohort, demographic and basic clinical data.

|  | Number of subjects | Percentage |
| --- | --- | --- |
| **Site** |  |  |
| Bonn | 38 | 44.7 |
| Cambridge | 5 | 5.9 |
| KCL | 32 | 37.6 |
| UCL | 10 | 11.8 |
| **Group** |  |  |
| HS | 23 | 27.1 |
| Healthy control | 39 | 45.9 |
| Disease control | 23 | 27.1 |
| **Sex** |  |  |
| Female | 36 | 42.4 |
| Male | 49 | 57.6 |
| **Age group** |  |  |
| Adult | 45 | 52.9 |
| Paediatric | 40 | 47.1 |
| **HS lateralisation (patients with HS)** |  |  |
| Left | 15 | 65.2 |
| Right | 4 | 17.4 |
| Bilateral | 4 | 17.4 |
| **Radiological MRI findings (patients with HS)** |  |  |
| 3T MRI negative, 7T MRI positive | 2 | 8.7 |
| 3T MRI negative, 7T MRI negative | 2 | 8.7 |
| 3T and 7T MRI positive | 19 | 82.6 |
| **Surgery (patients with HS)** |  |  |
| Resection | 8 | 34.8 |
| Ablation | 1 | 4.3 |
| None | 14 | 60.9 |
| **Confirmed by histopathology (patients with resections)** |  |  |
| Yes | 7 | 87.5 |
| Not available | 1 | 12.5 |
| **Favourable post-operative outcome (patients with any surgery)** |  |  |
| Yes (Engel/ILAE class I at 1 year) | 4 | 44.4 |
| No | 2 | 22.2 |
| Not available | 3 | 33.3 |

### 7T MRI acquisition parameters

7T T1-weighted images from the different sites were acquired using different sequences and parameters (Supplementary Table 1), which have been previously been described in detail [Dokumacı et al., 2023; Faber et al., 2026; Klodowski et al., 2025; Vecchiato et al., 2025]. 7T voxel sizes ranged between 0.6 and 0.8 mm isotropic. Data from Bonn were acquired using an MPRAGE sequence; for the other sites, MP2RAGE was used, which largely compensates for radiofrequency transmit field inhomogeneities [Marques et al., 2010]. For data from the UCL and KCL sites, T1-weighted ‘uniform’ MP2RAGE volumes had been preprocessed to generate ‘robust’ MP2RAGE, which removes salt-and-pepper background noise but can also re-introduce some bias field inhomogeneities [O’Brien et al., 2014].

### AID-HS predictions

HippUnfold segmentations within the AID-HS pipeline completed without manual intervention for all subjects. 4.1% (7/170) of reconstructions had a segmentation QC score below 0.7 (minimum 0.6); 5.9% (4/85) for 3T data and 3.5% (3/85) for 7T data. Manually reviewing these outputs, we found no clear segmentation errors and therefore the data were included in the subsequent analysis.

There were 19 patients with unilateral HS in the dataset. AID-HS detected, i.e., correctly predicted the presence and side of pathology, 12 HS using 3T data (sensitivity 63%) and 13 using 7T MR images (68%) (Table 2). The difference was not statistically significant (McNemar exact two-sided test p=1.0). The same 12 cases were correctly detected at both field strengths. A breakdown of detection sensitivity at 7T according to parallel transmit mode, whether denoising was applied to 7T T1w data, site and laterality of HS is provided in Supplementary Table 3. When assessing binary predictions of HS laterality (right/left, without permitting a ‘normal’ classification), 18 out of 19 HS (95%) were correctly lateralised using 3T data, and all 19 out of 19 when using 7T data.

**Table 2:** AID-HS classifier detection performance using 7T MRI data compared to paired 3T data. Differences between 3T and 7T performance are not statistically significant (McNemar p > 0.05) for all patients and controls or were not computed (for n=4 radiologically 3T MRI negative, n=2 radiologically 3T and 7T MRI negative, and n=7 histologically confirmed cases). These results were obtained without ComBat harmonisation of features. In a subset of the cohort where harmonisation was possible, results were not qualitatively different (Supplementary Table 2).

|  | 3T | 7T |
| --- | --- | --- |
| **All patients with unilateral HS** |  |  |
| Sensitivity for detection | 63.2% (12/19) | 68.4% (13/19) |
| Sensitivity for lateralisation | 94.7% (18/19) | 100.0% (19/19) |
| **Radiologically 3T MRI negative HS** |  |  |
| Sensitivity for detection | 50.0% (2/4) | 75.0% (3/4) |
| Sensitivity for lateralisation | 75.0% (3/4) | 100.0% (4/4) |
| **Radiologically 3T and 7T MRI negative HS** |  |  |
| sensitivity for detection | 50.0% (1/2) | 50.0% (1/2) |
| sensitivity for lateralization | 50.0% (1/2) | 100.0% (2/2) |
| **Histologically confirmed HS** |  |  |
| Sensitivity for detection | 57.1% (4/7) | 71.4% (5/7) |
| Sensitivity for lateralisation | 85.7% (6/7) | 100.0% (7/7) |
| **All controls** |  |  |
| Specificity | 96.8% (60/62) | 100.0% (62/62) |
| **Healthy controls** |  |  |
| Specificity | 97.4% (38/39) | 100.0% (39/39) |
| **Disease controls (FCD)** |  |  |
| Specificity | 95.7% (22/23) | 100.0% (23/23) |

Among the four radiologically 3T MRI-negative patients, two were detected by AID-HS at 3T and 7T. One additional MRI-negative patient was detected by AID-HS with 7T data. Given the small number of subjects, we did not compute a statistical hypothesis test.

Out of a total of 62 healthy and disease controls, 60 at 3T and 62 at 7T were correctly classified as having no asymmetry of hippocampal features suspicious of HS (difference p > 0.05). Specificity was high in both healthy and disease control subgroups. The only disease control misclassified at 3T (as right HS) had a suspected FCD in the right parahippocampal gyrus. At both field strengths, there were more controls lateralised towards the left (71.0%, 44/62), which was not the case when flipping inputs along the right/left axis (Supplementary Table 4). Flipping left and right sided inputs did not significantly affect detection performance (Supplementary Table 4).

Using T2-weighted images as inputs did not improve sensitivities but lowered specificity (Supplementary Table 6) and segmentation quality (Supplementary Figure 4).

### Example cases

See Figure 2 for two example cases with paired 7T and 3T images, hippocampal segmentations, extracted features and prediction scores. Case 1 (Patient 14 in [Klodowski et al., 2025]) had a prediction of HS only at 7T. This was in agreement with clinical findings, where HS had not been reported in the 3T but only in the 7T images. Case 2 represents a radiologically 3T and 7T MRI-negative HS case correctly predicted by AID-HS at both field strengths. The examples illustrate that the HippUnfold segmentation works reliably on MP2RAGE 7T data (Case 1) and on scans with signal dropout in the inferior temporal lobes (Case 2).

**Figure 2:**
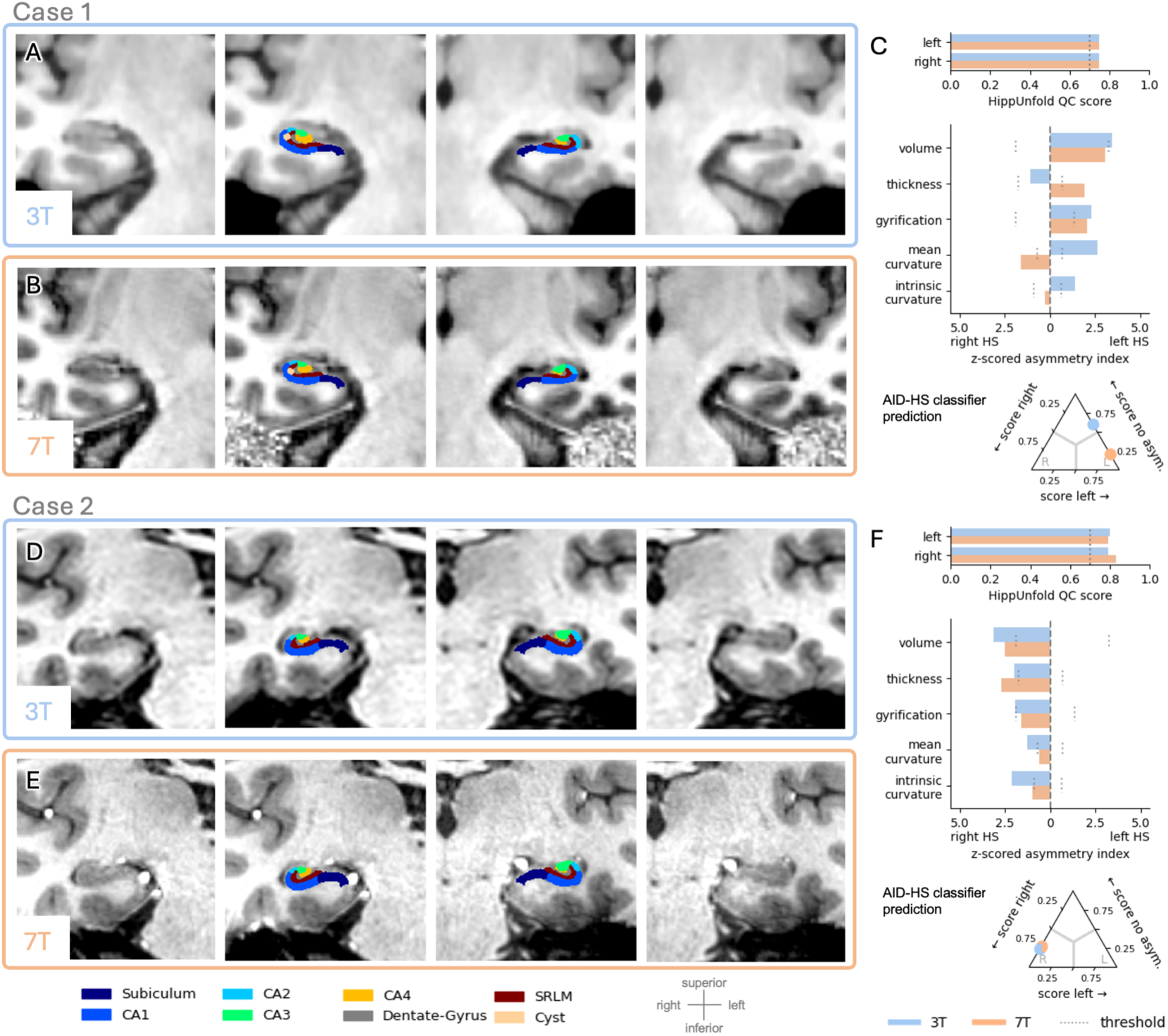
Example HippUnfold segmentations, AID-HS features and prediction scores compared between 7T and 3T data. Case 1 (A-C): male patient between age 40 and 45, 3T MRI-negative, radiological diagnosis of HS on 7T MRI, left temporal resection with histopathological confirmation of HS. Case 2 (D-F): female patient between age 30 and 35 with 3T and 7T MRI radiologically reported as unremarkable. Right selective amygdalo-hippocampectomy was performed and histology showed HS. Coronal slices of 3T MRI (A, D) and 7T MRI (B, E) of right and left hippocampus and overlaid high-resolution (0.2 mm) HippUnfold segmentations. 3T data was linearly co-registered and aligned to the 7T data for this visualisation. (C, F) comparison of HippUnfold segmentation QC score (with recommended threshold of 0.7 as dotted line), z-score normalized asymmetry features (with per-feature thresholds as dotted lines as in the original publication) and AID-HS prediction scores in ternary plots. For Case 1, segmentations are visually accurate for 3T and 7T data and quality control scores are identical. The AID-HS classifier prediction is “no asymmetry” for the 3T data and “left HS” for the 7T data. The thickness estimate from 7T data in this case is more consistent with a left sided HS. For case 2, hippocampal features extracted from data at both field strengths are similar and result in a high-confidence prediction of right HS in both cases. Note the MP2RAGE extra-cerebral noise (Case 1) and signal dropout in the lower temporal lobes (Case 2) do not affect the HippUnfold segmentation.

### Bilateral Hippocampal Sclerosis

There were a total of four patients with bilateral HS in the combined cohort. AID-HS enables comparison of feature values (e.g., hippocampal volume or thickness) to a normative control cohort. At both 3T and 7T, 3 of 4 bilateral HS cases were identified by three or more outlier features in both hippocampi and automatically flagged as having features consistent with bilateral HS. In healthy controls, this criterion had a specificity of 95.2% (59/62) at 3T and 93.5% (58/62) at 7T (difference not significant, McNemar test p>0.05). Harmonisation of features did not significantly change the performance (Supplementary Table 5). For the one out of four bilateral HS cases not identified by this criterion, classifier predictions and outlier features pointed towards a right hippocampal sclerosis; upon review of the clinical data, we noted this case had also initially been reported as a right HS. See Figure 3 for a bilateral HS example case. Of note, despite flagging 3 out of 4 cases as having features consistent with bilateral HS, the logistic regression classifier, which is designed to predict the side of unilateral HS or no asymmetry of hippocampal features, in these bilateral cases predicted left HS (1), right HS (2) or no asymmetry (1) both using 3T and 7T data.

**Figure 3:**
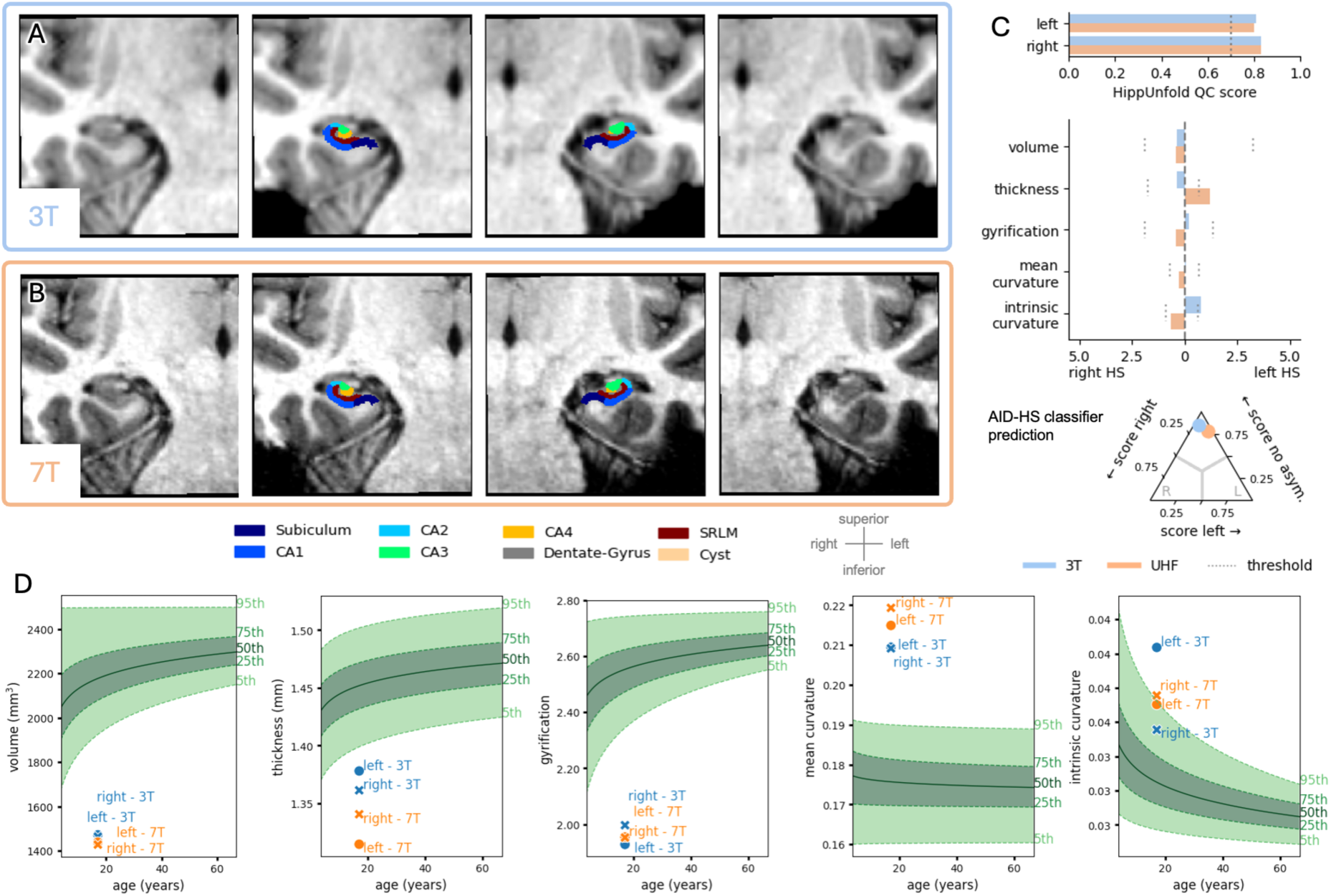
Example case with bilateral hippocampal sclerosis: male patient between age 15 and 20 with a clinical and radiological diagnosis of bilateral HS, not operated. Coronal slices of 3T (A) and 7T MRI (B) of right and left hippocampus and overlaid high-resolution (0.2 mm) HippUnfold segmentations. 3T data was linearly co-registered and aligned to the 7T data for this visualisation. The apparent resolution of 3T data is slightly reduced by interpolation but 7T slices are plotted without resampling. (C) Comparison of HippUnfold segmentation QC score (with recommended threshold of 0.7 as dotted line), z-score normalized asymmetry features (with per-feature thresholds as dotted lines as in the original publication) and AID-HS prediction score. (D) Hippocampal features plotted relative to generalized additive model percentiles of a reference cohort of healthy controls. Although the logistic regression classifier prediction is “no asymmetry”,4-5 of 5 features are above/below the 5th/95th percentile of the normative distribution, consistent with bilateral HS.

### Hippocampal features and prediction scores

We compared paired quantitative hippocampal measurements and prediction scores obtained using 3T and 7T MRI data (Figure 4). Among all hippocampal features extracted by AID-HS, we found a significant difference between field strengths for volume and thickness: In a mixed effects linear model, there was an average increase of volume of 50.0 mm^3^ (p_corr FDR_=0.012), and a decrease of thickness of 0.018 mm at 7T relative to 3T (p < 0.001) (Figure 4B). For the per-subject asymmetry scores, notably also volume and thickness asymmetries, (Figure 4F-J) and for the AID-HS logistic regression model prediction score (Figure 4K), we did not observe significant differences between field strengths. Intraclass correlation coefficients were: 0.82 (volume), 0.85 (thickness), 0.98 (gyrification), 0.77 (mean curvature), 0.92 (intrinsic curvature); 0.97, 0.83, 0.98, 0.70, 0.86 (asymmetries for the respective features); and 0.97 (AID-HS prediction score).

**Figure 4:**
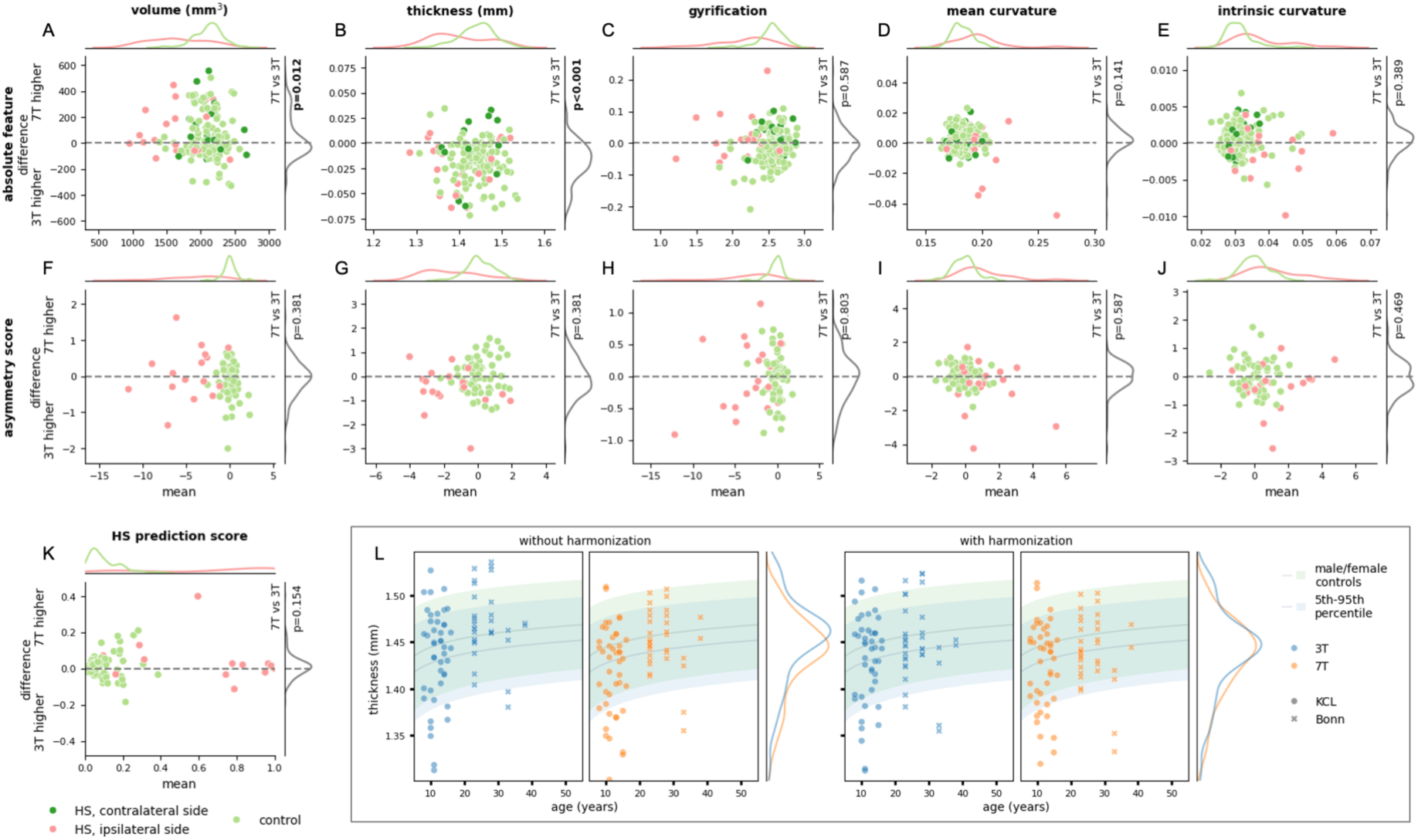
Quantitative comparison of hippocampal features estimated using 3T and 7T data, without ComBat harmonisation. (A-K) Bland-Altman plots (with difference/mean kernel density estimate plots along both axes) of: (A-E) the absolute hippocampal features extracted by HippUnfold and AID-HS (volume, thickness, gyrification index, mean curvature, intrinsic curvature) and (F-J) asymmetry scores of the same features; (K) AID-HS unilateral HS prediction score. Data from HS patients are coloured in red and dark green for ipsilateral and contralateral side, respectively, healthy and disease controls coloured in light green. For asymmetry scores in controls, a side was randomly chosen. For the effect of field strength (3T vs. 7T) on features in a mixed-effects linear regression model, t-test p-values with Benjamini-Hochberg FDR correction are stated (in bold if p<0.05). (L) Visualisation of the effect of ComBat harmonisation on hippocampal thickness for healthy controls. Scatter plots of thickness vs. age for 3T (blue) and 7T data (orange), without (left) and with harmonisation (right). Normative curves (AID-HS general additive model) 5th, 95th percentile and median are overlaid. At 7T, hippocampal volume is estimated slightly higher and thickness lower than at 3T. Asymmetry scores are not significantly different. Harmonisation may partly compensate for this effect see Supplementary Figure 2 for Bland-Altmann plots with ComBat-harmonized features.

For two sites, paired healthy controls were available (Bonn: 17, KCL: 22). We performed ComBat harmonisation using this data, which compensated for some of the observed differences in features across field strengths. Figure 4L illustrates that the healthy control data used for reference followed more similar distributions at 3T and 7T after harmonisation and aligned with the normative general additive model (GAM) used by AID-HS to facilitate the interpretation of quantitative features. However, with ComBat harmonisation, there were still significant differences between 3T and 7T features (Supplementary Figure 2).

Volume asymmetry indices derived with SynthSeg compared to HippUnfold yielded similar diagnostic performances (Supplementary Figure 3).

## Discussion

We collated a multi-centre, paired 3T and 7T MRI dataset for validation of the research software tool AID-HS for automated detection of hippocampal sclerosis at 7T. We found no evidence of differences in sensitivity of AID-HS for detecting and lateralizing HS (60-70% and close to 100%, respectively) and high specificity in controls (above 95%) across field strengths. AID-HS correctly identified two out of four radiologically 3T MRI-negative cases using 3T MRI data, and one additional case when using 7T MRI data. Overall, the features and asymmetry indices showed high agreement between 7T and 3T MRI, with hippocampal volume being on average higher and thickness slightly lower at 7T. For flagging bilateral HS, using the criterion of three or more (out of five) outlier features in both hippocampi correctly identified three out of four such cases in our cohort (i.e., sensitivity ∼75%), while specificity in controls was around 95% - at both field strengths.

AID-HS performance was robust to the range of generalisation challenges which computational neuroimaging software faces with 7T MRI. These include higher resolution images and increased radiofrequency transmit field inhomogeneity. The resulting image artefacts are often located in the temporal lobes, and might explain the increased segmentation errors found for other softwares at 7T [Burkett et al., 2025]. In this cohort, such artefacts were visible in many of the 7T images, but Hippunfold segmentation and AID-HS performance was robust to these effects (see Figure 2 D-F for an example case). We observed on average higher volume and lower thickness estimates at 7T, but generally, hippocampal features showed a high agreement between field strengths. The effects of 7T acquisitions on hippocampal volume measurements appear to vary for different segmentation software [Burkett et al., 2025; Seiger et al., 2015], which may be due to different contributions of T1, T2* and PD contrast to 7T images [Lorio et al., 2016]. In this dataset, hippocampal volumes derived with SynthSeg – as a widely used segmentation tool that is designed to be robust across different contrasts and modalities [Billot et al., 2023] – yielded downstream performance for classification of HS that was comparable to HippUnfold volumes, at both field strengths (Supplementary Figure 3). The AID-HS classifier uses additional morphometric features beyond volume that can provide more diagnostic information (thickness, gyrification, curvature). The combination of state-of-the-art feature extraction, normalisation (by per-subject asymmetry scoring as well as ComBat harmonisation) and simple, interpretable classifier design likely all contribute to ensuring that AID-HS is robust to domain shift from 3T to 7T MRI data.

The sensitivity for HS detection at both 3T and 7T was lower in this study than that reported in the original AID-HS testing cohort (∼90% overall, ∼80% in MRI-negative HS) [Ripart et al., 2024a]. This could be due to greater uncertainty with a smaller cohort (n=19 vs n=256) and potentially also due to selection bias. Cases recruited or referred for 7T MRI by clinicians are likely to be diagnostically more difficult. Indeed, for the Cambridge cohort, definitive findings at 3T were an explicit exclusion criterion. Although only four patients in our combined cohort were truly radiologically 3T MRI negative, this designation reflects repeated 3T imaging and careful clinical review with a low threshold for reporting subtle abnormalities. It should also be noted that radiological reports served as ground truth in most cases when histopathology was not available, which makes evaluation of detection performance less certain. Nevertheless, lateralisation sensitivity of AID-HS was 94.7% for 3T and 100% 7T. Volumetric hippocampal atrophy was less pronounced in HS patients missed by AID-HS compared to those correctly predicted (Supplementary Figure 1), suggesting that failures occurred mainly in the most subtle cases.

Among the patients who were 3T MRI negative, 7T MRI had allowed a visual radiological diagnosis of HS in two out of four cases. This is consistent with different studies describing an increased yield of 7T MRI for detecting hippocampal abnormalities [Burkett et al., 2026; Hangel et al., 2024; van Lanen et al., 2021]. AID-HS correctly predicted HS in two of these four using 3T data only, and in three when using 7T MRI data. Although the low number of cases limits statistically valid conclusions, this illustrates how AID-HS might be clinically useful in difficult to diagnose patients.

Bilateral HS is particularly challenging to diagnose as radiological and computational approaches often rely on feature asymmetries with the contralateral hippocampus to guide the diagnosis. AID-HS reports automatically flagged the majority of bilateral HS cases as exhibiting outlier hippocampal features consistent with bilateral HS relative to normative thresholds at both 3T and 7T (Supplementary Table 5). This highlights the practical value of interpretable morphological features in the AID-HS pipeline, which complement classifier outputs in situations such as bilateral HS.

A slight predominance of left-sided HS has been reported (55%, [Whelan et al., 2018]). In this cohort, there is a more pronounced skew towards left-HS cases (79%). For the AID-HS classifier, there was evidence of a higher prior probability of left-sided lateralisation in healthy controls. However, when flipping left and right sides of inputs, diagnostic performance remained high (Supplementary Table 4).

A strength of this study is its multi-centre cohort that incorporates data from different sites with variability in scanners, acquisition protocols and patient populations (paediatric and adult), albeit with two sites contributing the majority of cases. External validation is an important step towards clinical translation of medical imaging software. Large multi-centre collaborations have allowed progress on clinically relevant issues for neuroimaging in epilepsy with 3T MRI [Gill et al., 2021; Ripart et al., 2024b; Wagstyl et al., 2022; Whelan et al., 2018]. While 7T MRI is still a relatively scarce investigation, and pathologies causing DRFE are heterogeneous or may remain uncertain in some cases, pooling data from multiple sites is the most feasible way to obtain reasonable numbers of cases for studying specific pathologies. Here, we were able to gather 23 HS cases from four sites. We believe that further collaborative efforts will be necessary to leverage the full potential of 7T MRI in DRFE with state-of-the-art computational methods.

It should be noted that we used 7T MRI T1-weighted sequences with a highest resolution of 0.6 mm isotropic voxels because these data were available for the greatest number of subjects and because AID-HS was developed for these inputs. High in-plane resolution coronal T2-weighted scans are considered the gold standard for assessing hippocampal pathology, and the consensus recommendations (ILAE HARNESS protocol) include a 0.4x0.4x2.0mm coronal T2w sequence at 3T [Bernasconi et al., 2019]. Higher resolution 7T images can improve the precision of hippocampal morphometry and delineation of hippocampal subfields [Henry et al., 2011; Stefanits et al., 2017]. This is of particular interest to allow assignment to pathological subtypes [Blümcke et al., 2013] and potentially to improve sensitivity in subtle cases where overall hippocampal volume may be relatively preserved, as in subtypes 2 and 3 [Middlebrooks et al., 2024]. Subfield parcellation is provided by HippUnfold (Figures 1, 2) but currently not used by AID-HS, which averages features across the whole hippocampus, favouring model simplicity and interpretability. HippUnfold-estimated subfield volumes at 3T have been shown to distinguish histopathological subtypes [Hung et al., 2026], suggesting that this pipeline could be extended, although the present dataset does not comprise histopathological data or a sufficient number of cases to train a dedicated classifier. Applying AID-HS, which was developed for T1-weighted inputs, on T2-weighted images did not improve segmentation quality or diagnostic performance in this study (Supplementary Table 6, Supplementary Figure 4). Future studies combining multicentre, multimodal 7T imaging, quantitative subfield analysis and detailed histopathological subtyping may result in automated pipelines with further increases in sensitivity and the ability to perform HS subtyping.

The main limitation of this work is the small sample size, especially concerning 3T MRI negative patients and bilateral HS, as well as the lack of histological confirmation of HS in many patients. This is in part due to the current scarcity of 7T scanners as well as the overall likelihood that HS will be detected on conventional (3T) MRI and not subsequently imaged at 7T. As MRI examinations at some sites were recent, some patients in this cohort will still proceed to epilepsy surgery and follow-up clinical data can be expected to become available. Overall, because the number of patients with verified HS and normal 3T scans who have undergone 7T MRI remains very small, the statistical power to test for possible superiority of 7T MRI detection performance is limited.

In conclusion, AID-HS provides consistent quantitative estimates of hippocampal features and reliable predictions for detecting and lateralizing HS in 7T MRI, despite having been developed with 3T MRI data. This facilitates the further integration of 7T MRI in the clinical management of DRFE. Realising potential further diagnostic gains of 7T MRI with automated detection of HS will likely require larger datasets and dedicated models. These results motivate investigating how other software tools for the detection of epileptogenic lesions such as FCDs [Bernasconi et al., 2019; Kersting et al.,; Ripart et al., 2024b; Ripart et al., 2025] perform on 7T MRI data.

## Data Availability

The MRI data cannot be shared publicly due to privacy concerns. The code used for analysis is available in a github repository (https://github.com/ckronlage/aidhs_uhf_pub/).

## Acknowledgements

CK is supported by the DFG and the Faculty of Medicine Tuebingen (MINT-CS), the Else Kroener Fresenius Stiftung (PRECISE.net), the Michael Foundation, de.NBI Cloud within the German Network for Bioinformatics Infrastructure (de.NBI) and ELIXIR-DE. MR and KW are supported by the Wellcome Trust (301991/Z/23/Z). RJP is supported by GOSH Children’s Charity. TB is supported by GOSH Biomedical Research Centre (BRC). JSD is supported by NIHR. MHE is supported by The Foulkes Foundation. TB is supported by Neuro-aCSis Bonn Neuroscience Clinician Scientist Programme (German Research Foundation, Deutsche Forschungsgemeinschaft, DFG, 493623632). TR is supported by the German Ministry for Research, Technology and Space (epi-center.ai). KK, CTR, TEC are supported by the Medical Research Council: Developmental pathway funding scheme UKRI790, Mid-range equipment fund award UKRI2679, University of Cambridge Impact Acceleration Account Award MR/X502844/1, Clinical Research Infrastructure Award MR/M008983/1, the Cambridge University Hospitals Academic Fund for functional neurosurgery, the NIHR Cambridge Biomedical Research Centre (NIHR203312). SA is supported by Vera Down Grant, BMA Foundation. This article represents independent research in part funded by the infrastructure of the National Institute for Health Research (NIHR) Mental Health Biomedical Research Centre (BRC) at South London and Maudsley NHS Foundation Trust and King’s College London. This research was funded/supported by the National Institute for Health and Care Research (NIHR) Clinical Research Facility at Guy’s and St Thomas’ NHS Foundation Trust. The content of this manuscript is solely the responsibility of the authors, the views expressed are those of the authors and not necessarily those of the NHS, the NIHR, the Department of Health and Social Care or any of the other funders.The funders had no role in study design, data collection and analysis, decision to publish or preparation of the manuscript.

