## Supplementary Material for "Automated hippocampal sclerosis detection, using AID-HS, shows robust performance across multi-centre paired 7T and 3T MRI"

| $B_0$ | Scanner | Sequence | pTx mode | Voxel dimensions |
|---|---|---|---|---|
| **Bonn** | | | | |
| 7T | Siemens Magnetom 7T Plus | MPRAGE | UP | 0.6 mm isotropic |
| 3T | 2 different scanners (Philips Achieva, Siemens Tim Trio) | different 3D T1w | | 0.5/0.8/1.0 mm isotropic |
| **Cambridge** | | | | |
| 7T | Siemens Magnetom Terra | MP2RAGE | CP | 0.8 mm isotropic |
| 3T | 2 different scanners (GE Discovery MR750, GE Signa Premier) | different 3D T1w | | 0.5/1.0 mm isotropic |
| **KCL** | | | | |
| 7T | Siemens Magnetom Terra | MP2RAGE | CP | 0.56/0.60/0.65 mm isotropic |
| 3T | Philips Achieva 3T | MPRAGE | | 1.0 mm isotropic |
| **UCL** | | | | |
| 7T | Siemens Magnetom Terra | MP2RAGE | CP | 0.65 mm isotropic |
| 3T | 3 different scanners (GE Discovery MR750, Siemens Magnetom Vida, Siemens Magnetom Prisma) | different 3D T1w | | 1.0 mm isotropic |

Supplementary Table 1: Overview of MRI systems and acquisition parameters of data acquired at different sites. $B_0$: static magnetic field strength. MPRAGE: Magnetization Prepared Rapid Gradient Echo. MP2RAGE: Magnetization Prepared 2 Rapid Gradient Echo. T1w: T1-weighted. pTx mode: All scanners used head coils with multiple transmit elements, either with universal pulses (UP) or in circularly polarized (CP) mode.

| | No harmonisation | | With harmonisation | |
|---|---|---|---|---|
| | **3T** | **7T** | **3T** | **7T** |
| **Patients with unilateral HS** | | | | |
| Sensitivity for detection | 81.8% (9/11) | 81.8% (9/11) | 81.8% (9/11) | 81.8% (9/11) |
| Sensitivity for lateralisation | 100.0% (11/11) | 100.0% (11/11) | 100.0% (11/11) | 100.0% (11/11) |
| **Radiologically 3T MRI negative HS patients** | | | | |
| Sensitivity for detection | 100.0% (1/1) | 100.0% (1/1) | 100.0% (1/1) | 100.0% (1/1) |
| Sensitivity for lateralisation | 100.0% (1/1) | 100.0% (1/1) | 100.0% (1/1) | 100.0% (1/1) |
| **Radiologically 3T and 7T MRI negative HS patients** | | | | |
| Sensitivity for detection | 100.0% (1/1) | 100.0% (1/1) | 100.0% (1/1) | 100.0% (1/1) |
| Sensitivity for lateralisation | 100.0% (1/1) | 100.0% (1/1) | 100.0% (1/1) | 100.0% (1/1) |
| **Histologically confirmed HS** | | | | |
| Sensitivity for detection | 100.0% (2/2) | 100.0% (2/2) | 100.0% (2/2) | 100.0% (2/2) |
| Sensitivity for lateralisation | 100.0% (2/2) | 100.0% (2/2) | 100.0% (2/2) | 100.0% (2/2) |
| **All controls** | | | | |
| Specificity | 96.5% (55/57) | 100.0% (57/57) | 96.5% (55/57) | 100.0% (57/57) |
| **Healthy controls** | | | | |
| Specificity | 97.4% (38/39) | 100.0% (39/39) | 97.4% (38/39) | 100.0% (39/39) |
| **Disease controls (FCD)** | | | | |
| Specificity | 94.4% (17/18) | 100.0% (18/18) | 94.4% (17/18) | 100.0% (18/18) |

Supplementary Table 2: AID-HS classifier detection performance with ComBat harmonisation for cohorts where healthy controls were available.

| | Sensitivity for detection |
|---|---|
| All subjects | 68.4% (13/19) |
| **pTx mode** | |
| Universal pulses (UP) | 80.0% (8/10) |
| Circularly polarized (CP) | 55.6% (5/9) |
| **Type of 7T T1w sequence** | |
| MP2RAGE with background noise | 50.0% (1/2) |
| MEMPRAGE or denoised MP2RAGE | 70.6% (12/17) |
| **Site** | |
| Bonn | 80.0% (8/10) |
| Cambridge | 50.0% (1/2) |
| KCL | 100.0% (1/1) |
| UCL | 50.0% (3/6) |
| **Laterality of HS** | |
| Right | 75.0% (3/4) |
| Left | 66.7% (10/15) |

Supplementary Table 3: AID-HS detection sensitivity using 7T T1w data in different subgroups. There was no significant difference in sensitivity to detect HS in each subgroup (Fisher's exact test for pTx mode, type of T1w sequence and laterality of HS, permutation Chi-square test for comparison of the sites). However, statistical power is limited due to the small sample size.

| | 3T | | 7T | |
|---|---|---|---|---|
| | original | right-left flipped | original | right-left flipped |
| **Patients with unilateral HS** | | | | |
| Sensitivity for detection | 63.2% (12/19) | 68.4% (13/19) | 68.4% (13/19) | 68.4% (13/19) |
| Sensitivity for lateralisation | 94.7% (18/19) | 89.5% (17/19) | 100.0% (19/19) | 84.2% (16/19) |
| **Radiologically 3T MRI negative HS patients** | | | | |
| Sensitivity for detection | 50.0% (2/4) | 75.0% (3/4) | 75.0% (3/4) | 75.0% (3/4) |
| Sensitivity for lateralisation | 75.0% (3/4) | 75.0% (3/4) | 100.0% (4/4) | 75.0% (3/4) |
| **Radiologically 3T and 7T MRI negative HS patients** | | | | |
| Sensitivity for detection | 50.0% (1/2) | 50.0% (1/2) | 50.0% (1/2) | 50.0% (1/2) |
| Sensitivity for lateralisation | 50.0% (1/2) | 50.0% (1/2) | 100.0% (2/2) | 50.0% (1/2) |
| **Histologically confirmed HS** | | | | |
| Sensitivity for detection | 57.1% (4/7) | 71.4% (5/7) | 71.4% (5/7) | 71.4% (5/7) |
| Sensitivity for lateralisation | 85.7% (6/7) | 85.7% (6/7) | 100.0% (7/7) | 71.4% (5/7) |
| **All controls** | | | | |
| Specificity | 96.8% (60/62) | 96.8% (60/62) | 100.0% (62/62) | 96.8% (60/62) |
| **Healthy controls** | | | | |
| Specificity | 97.4% (38/39) | 97.4% (38/39) | 100.0% (39/39) | 97.4% (38/39) |
| **Disease controls (FCD)** | | | | |
| Specificity | 95.7% (22/23) | 95.7% (22/23) | 100.0% (23/23) | 95.7% (22/23) |

Supplementary Table 4: AID-HS classifier detection performance with original input data and inputs flipped along the right-left axis. Classifier outputs concerning lateralisation in controls (i.e., the side of larger HS prediction score, irrespective of overall prediction) showed a bias toward the left with 71.0% (44/62) left lateralised vs. 29.0% (18/62) right lateralised, at both 3T and 7T (binomial test $p < 0.05$ for a null hypothesis of $P=0.5$). In the right-left flipped conditions there were 51.6% (32/62) at 3T and 48.4% (30/62) left predictions ($p > 0.5$).

| | No harmonisation | | With harmonisation | |
|---|---|---|---|---|
| | **3T** | **7T** | **3T** | **7T** |
| **All cohorts** | | | | |
| Sensitivity (bilateral HS) | 75.0% (3/4) | 75.0% (3/4) | - | - |
| Specificity (controls) | 95.2% (59/62) | 93.5% (58/62) | - | - |
| **Cohorts with healthy controls available** | | | | |
| Sensitivity (bilateral HS) | 50.0% (1/2) | 50.0% (1/2) | 50.0% (1/2) | 50.0% (1/2) |
| Specificity (controls) | 98.2% (56/57) | 96.5% (55/57) | 98.2% (56/57) | 96.5% (55/57) |

Supplementary Table 5: Classification performance of bilateral HS based on outlier hippocampal features, without and with ComBat harmonisation.

| | 3T T1w | 3T T2w | 7T T1w | 7T T2w |
|---|---|---|---|---|
| **Patients with unilateral HS** | | | | |
| Sensitivity for detection | 81.8% (9/11) | 90.9% (10/11) | 81.8% (9/11) | 90.9% (10/11) |
| Sensitivity for lateralisation | 100.0% (11/11) | 81.8% (9/11) | 100.0% (11/11) | 81.8% (9/11) |
| **Radiologically 3T MRI negative HS patients** | | | | |
| Sensitivity for detection | 100.0% (1/1) | 0.0% (0/1) | 100.0% (1/1) | 100.0% (1/1) |
| Sensitivity for lateralisation | 100.0% (1/1) | 100.0% (1/1) | 100.0% (1/1) | 0.0% (0/1) |
| **Radiologically 3T and 7T MRI negative HS patients** | | | | |
| Sensitivity for detection | 100.0% (1/1) | 0.0% (0/1) | 100.0% (1/1) | 100.0% (1/1) |
| Sensitivity for lateralisation | 100.0% (1/1) | 100.0% (1/1) | 100.0% (1/1) | 0.0% (0/1) |
| **Histologically confirmed HS** | | | | |
| Sensitivity for detection | 100.0% (2/2) | 50.0% (1/2) | 100.0% (2/2) | 100.0% (2/2) |
| Sensitivity for lateralisation | 100.0% (2/2) | 100.0% (2/2) | 100.0% (2/2) | 50.0% (1/2) |
| **All controls** | | | | |
| Specificity | 92.3% (12/13) | 69.2% (9/13) | 100.0% (13/13) | 61.5% (8/13) |
| **Healthy controls** | | | | |
| Specificity | 100.0% (4/4) | 100.0% (4/4) | 100.0% (4/4) | 50.0% (2/4) |
| **Disease controls (FCD)** | | | | |
| Specificity | 88.9% (8/9) | 55.6% (5/9) | 100.0% (9/9) | 66.7% (6/9) |

Supplementary Table 6: AID-HS classifier detection performance with T2w inputs, compared to the default T1w inputs, for all subjects with both T1w and T2w available at 3T and 7T. Using T2-weighted data did not consistently improve sensitivity and lowered specificity. In-plane resolution of 7T T2-weighted images was 0.44 mm. HippUnfold segmentation quality control (QC) scores were significantly lower with T2-weighted images (Supplementary Figure 4).

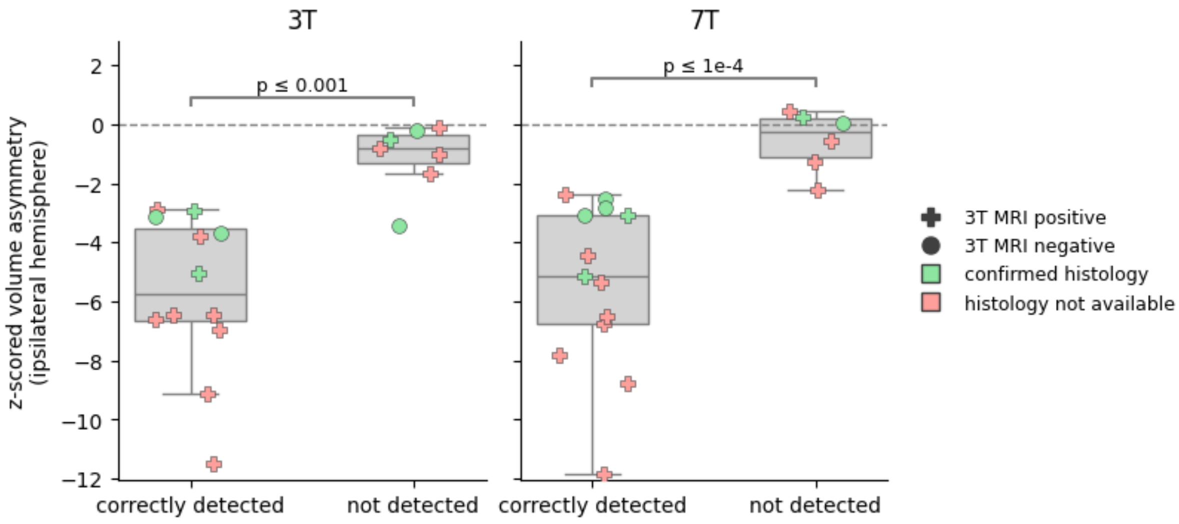


Supplementary Figure 1: For unilateral HS patients, ipsilateral volume asymmetry indices according to subgroups of patients correctly detected by AID-HS (left) vs. not detected (right). Boxplots and overlaid scatterplots, individual data points are shaped according to visual 3T MRI findings (positive/negative) and coloured to reflect histopathological data (green: confirmed, red: not available). Volume asymmetry scores in the subgroups are statistically different (Mann-Whitney-U test). HS patients correctly detected by AID-HS have more pronounced volumetric unilateral atrophy. A limitation for this analysis is that volume asymmetry scores and AID-HS predictions are not independent, as volume asymmetry is an input of the classifier.

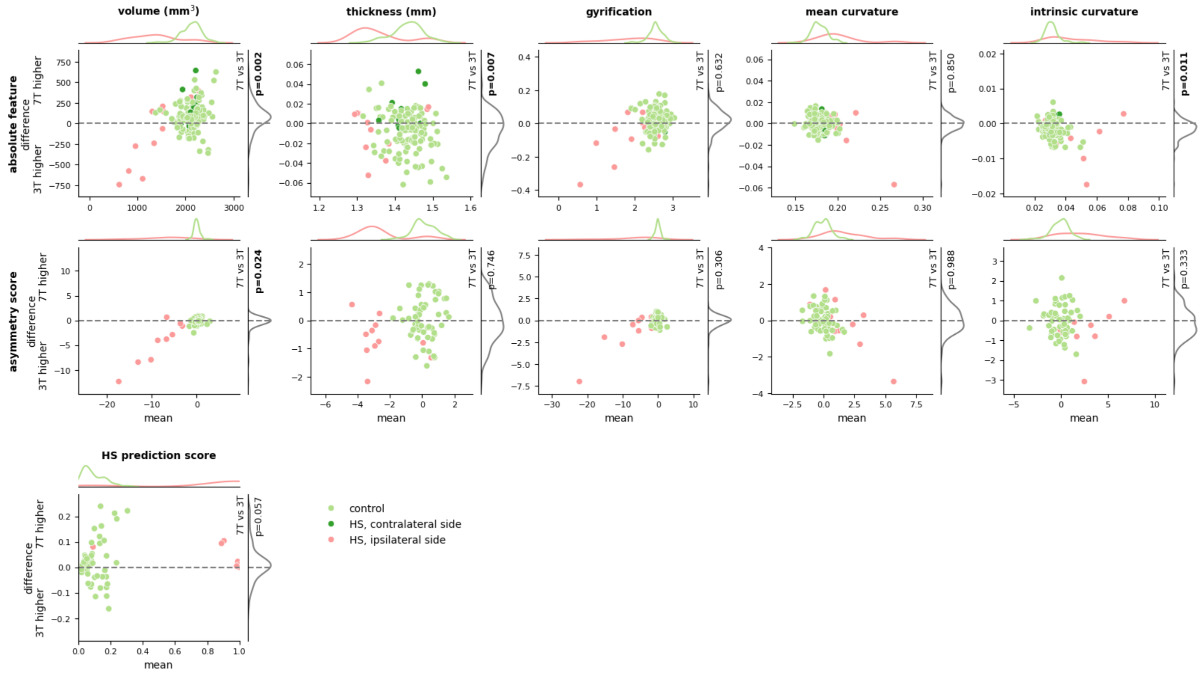


Supplementary Figure 2: Comparison of hippocampal features obtained at 7T vs. 3T with ComBat harmonisation for the two cohorts with healthy controls available (analogous to Figure 3). Bland-Altman plots (with kernel density estimate plots along both axes) of: (top row) the absolute hippocampal features extracted by HippUnfold and AID-HS (volume, thickness, gyrification index, mean curvature, intrinsic curvature); (second row) asymmetry scores of the same features; (third row) AID-HS unilateral HS prediction score. Data from HS patients are coloured in red and dark green for ipsilateral and contralateral side, respectively, healthy and disease controls coloured in light green. For asymmetry scores in controls, a side was randomly chosen. For the effect of field strength (3T vs. 7T) on features in a mixed-effects linear regression model, t-test p-values with Benjamini-Hochberg FDR correction are stated (in bold if significant $p<0.05$). With harmonisation, estimates for hippocampal thickness show reduced differences across field strengths, although these remained statistically significant. For other features (volume, gyrification, mean and intrinsic curvature), outlier values in HS appear more pronounced after harmonisation. This could be due to slightly lower variance in healthy controls at 7T, leading to estimation of larger site-specific scaling factors.

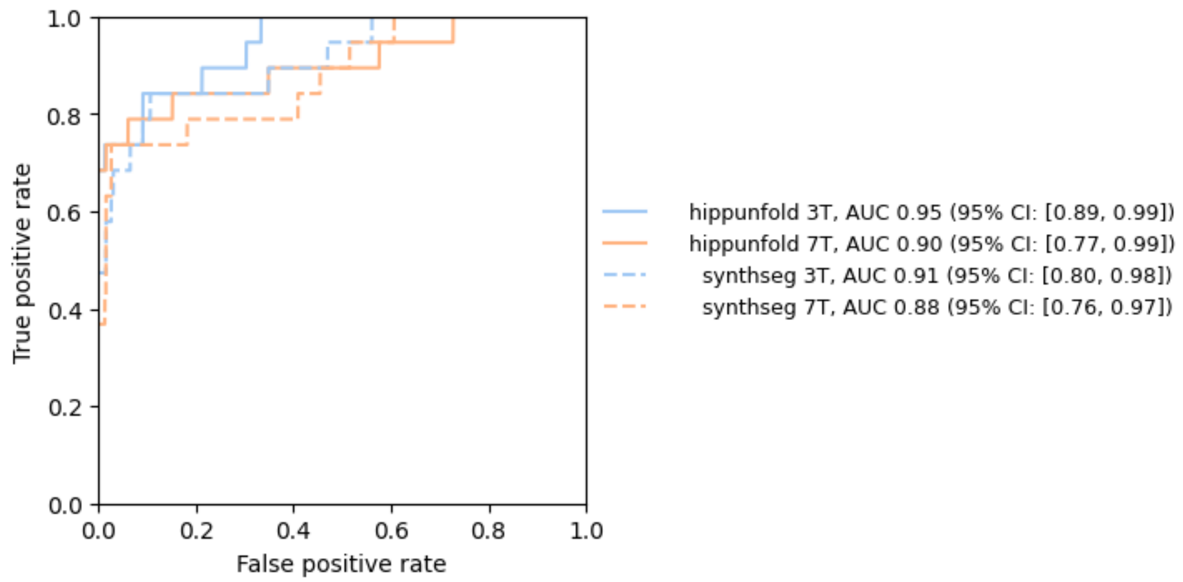


Supplementary Figure 3: Receiver operating characteristic analysis (ROC) analysis for the binary classification of subjects as right HS vs. no or left HS and left HS vs. no or right HS, based on hippocampal volume asymmetry indices, weighted average (by the number of left and right HS cases. AUC is the area under the ROC curve. 95% confidence intervals were obtained by bootstrapping (resampling with replacement) for 10,000 iterations. HippUnfold segmentation (continuous line) and SynthSeg segmentation (dashed line) for 3T (blue) and 7T (orange) data. Confidence intervals overlap. There is a trend for performance to be slightly higher for HippUnfold compared to SynthSeg and slightly higher for 3T data compared to 7T data.

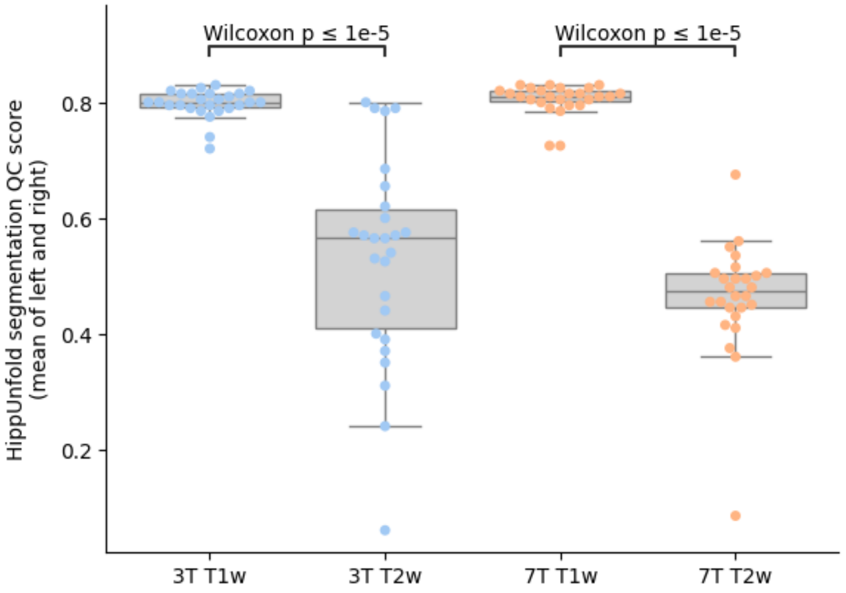


Supplementary Figure 4: HippUnfold segmentation quality control (QC) score, per-subject mean of left and right, in matched 3T and 7T T1-weighted and T2-weighted images. Segmentation QC scores are significantly lower when using T2-weighted data.
